# Air pollution exposure among people with limitations in activities of daily living in the United States

**DOI:** 10.64898/2025.12.22.25342861

**Authors:** Heather McBrien, Maddie Taylor, Marissa L. Childs, Lara Schwarz, Katherine Wolf, Marianthi-Anna Kioumourtzoglou, Rachel Morello-Frosch, Joan A. Casey

## Abstract

Structural barriers including limited healthcare access and disability-related health conditions make disabled people differentially susceptible to air pollution-related adverse health outcomes compared to nondisabled people. We used 2020 census-tract level counts of individuals with limitations in activities of daily living (ADLs) to identify a subset of disabled people. We described geographic areas where this population was highly exposed to air pollution in the contiguous U.S., indicating health risk.

We assessed census tract-level exposure to PM_2.5_, O_3_, NO_2_ (2016–2020), and wildfire PM_2.5_ (2016–2023). We mapped high ADL limitation prevalence and high air pollution exposure census tracts. Because environmental injustice means race and poverty strongly predict air pollution exposure, we also assessed exposure among people with ADL limitations by these demographic factors to identify doubly vulnerable subpopulations.

High ADL limitation prevalence and PM_2.5_/NO_2_ exposure co-occurred in urban areas, California’s Central Valley, Eastern Washington, and parts of the Southeast. Among people with ADL limitations, Asian and Hispanic individuals and those experiencing poverty were more exposed to PM_2.5_, O_3_, and NO_2_.

Disability is not fully captured by ADL limitations; future studies should explore other definitions of disability. Future studies should evaluate interventions to reduce air pollution-related morbidity and mortality, especially in regions and subpopulations identified here, where disabled people face high exposure and multiple vulnerabilities.

## Introduction

Disabled people have recognized the unique risks air pollution poses to their community and mobilized to reduce those risks.^1,2,3,4,5^ Yet, they remain at higher risk of air pollution-related disease onset, disease exacerbations, and mortality compared to nondisabled people.^6^ Structural ableism^7^ –barriers to healthcare, inaccessible risk communication and emergency response, and exclusion from education, employment, housing, and transportation systems–makes disabled people vulnerable to air pollution-related disease.^8,9^ Mobility limitations and disability-related health conditions may further increase risks.^6^

Increasingly prevalent wildfire smoke^10^ may be particularly harmful for disabled people. Inaccessible transportation may prevent disabled people from temporarily relocating away from smoke, medical devices may preclude masking to reduce exposure, and those receiving disability benefits may not have income available to purchase air filters.^11,6,7,12,13^ Urgent healthcare for smoke-related symptoms may be inaccessible.^12^ Nursing facilities or prisons housing disabled people may not respond to smoke emergencies adequately or at all,^14^ and unhoused disabled people may face extremely high outdoor exposures.

Despite this vulnerability, public health and environmental health researchers rarely study disability as a social factor influencing health.^15,16^ Rather, they treat disability as a physiological deficit or an undesirable adverse health outcome.^8,17,18^ In fact, a range of structural factors (structural ableism) beyond individual impairments restrict opportunities for disabled people and influence health outcomes. ^7,17,19^ These structural determinants create health disparities between disabled and nondisabled people.^19^

In 2023, the National Institutes of Health designated disabled people as a health disparity population for the first time,^16^ placing disability alongside race, gender, and socioeconomic status as a social factor influencing health. This designation called for research on the disproportionate impacts of environmental hazards on disabled people. This new understanding could allow public health research to shift from ‘curing’ disability (which disabled people have critiqued as rooted in eugenics)^7^ to analyzing structural causes of disability-related disparities.

One previous study has examined air pollution exposure among disabled people while treating disability as a social factor influencing health.^20^ Chakraborty found that higher average census-tract scale PM_2.5_ exposure 2011-2015 was associated with a higher percentage of disabled people in U.S. census tracts. This association persisted after controlling for other factors: race and ethnicity, poverty, renter occupancy, age, population density, and metropolitan status.^20^ These results suggest disability may be associated with higher air pollution exposure. However, they do not describe where disabled people are highly exposed to air pollution, or whether disabled people are more exposed to air pollution compared to nondisabled people. Also, Chakraborty did not include people in institutions or those without a residential address. Because disability discrimination often involves institutionalization,^21^ incarceration,^22,23^ or impoverishment,^24^ including these populations is essential.

Here, we adopt a critical disability studies framework, understanding disability as a social factor influencing vulnerability to air pollution-related health effects, alongside race and socioeconomic status. We describe where disabled people in the contiguous U.S. are highly exposed to air pollution, indicating health risk. We used American Community Survey (ACS) data to estimate the number of people reporting limitations in activities of daily living (ADLs) in each census tract, approximating a subset of disabled people. We assessed this population’s exposure to four air pollutants: fine particulate matter (PM_2.5_), ozone (O_3_), nitrogen dioxide (NO_2_) (2016-2020), and wildfire PM_2.5_ (2016-2023). We mapped areas with both high prevalence of ADL limitations and high air pollution exposure. We also identified populations facing dual burdens of social and environmental exposure by assessing exposure among people with ADL limitations by race and poverty status.

## Methods

### Study population and demographic variables

We used 2016-2020 ACS data to estimate the number of people reporting limitations in ADLs in each census tract, approximating a subset of disabled people. ACS respondents could report difficulties in hearing, vision, cognitive, ambulatory, self-care, or independent living.^25–27^ These estimates did not include ADL limitation prevalence for people living in “group quarters” including jails, prisons, shelters, and nursing facilities, or for unhoused people at the census tract scale.^27^

To estimate the number of people living in group quarters in each census tract, we used 2020 Census data. To estimate ADL limitation prevalence in group quarters, we used both published estimates and estimates of the national prevalence of ADL limitations in group quarters from the 2016–2020 ACS (which did not include census tract scale estimates). We used ADL limitation prevalences of 40% among adults in prisons and jails,^28–31^ 60% among incarcerated children,^32,33^ 60% among shelter residents^34,35^, and 100% among nursing facility residents.^34^ We used ADL limitation prevalences of 60% for individuals in psychiatric institutions and 12% for those in college dorms and military barracks, from national ACS estimates only,^34^ since no other published estimates were available. We were unable to include unhoused individuals outside shelters due to lack of census tract scale data.

We used ACS data to estimate census tract scale counts of people with ADL limitations by race and ethnicity (non-Hispanic Black, White, Asian American, Pacific Islander or Native Hawaiian, American Indian and Alaska Native, Other, and Two or more races, as well as Hispanic), poverty status (those with income in the last twelve months below the 2020 federal poverty threshold income versus at or above poverty threshold), and age (<18, 18-64, and 65+). These data were not available for people in group quarters.

We used 2010 revised Rural-Urban Commuting Area (RUCA) codes (the most recent as of publication) to classify tracts as Metropolitan (Urban), Micropolitan (Large Town), Small Town, or Rural (Isolated Rural). We categorized Metropolitan tracts as urban, and all others as rural.^36^

We included population counts from contiguous U.S. tracts in the Census Bureau’s 2010 TIGER/Line Files only, since air pollution estimates with comparable predictive accuracy were unavailable for tracts outside the contiguous U.S.

This study relied on publicly available de-identified data and thus did not constitute human subjects research.

### Exposure assessment

We estimated long-term exposure to four ambient air pollutants (PM_2.5_, O_3_, NO_2_, and wildfire PM_2.5_), chosen because they have been consistently associated with adverse health outcomes, ^37–39^ and worse adverse health outcomes in disabled people versus nondisabled people.^40^ We assessed exposure for 2010 census tracts and cross-walked exposure estimates to 2020 census tracts using the United States Census Bureau Census Tract Relationship File. We assigned exposures by 2020 census tract of residence.^41^

We used predictions from the Community Multiscale Air Quality (CMAQ) Fused Air Quality Surface Using Downscaling model to assess PM_2.5_ and O_3_ exposure.^42^ CMAQ estimates are publicly available, span recent years (2016-2020), and were specifically designed in the fused model to predict pollutant concentrations for 2010 census tracts. We averaged daily predicted mean outdoor PM_2.5_ concentrations at tract centroids 2016–2020. For O_3_, we averaged predicted 8-hour daily maximum ozone concentrations at each tract centroid over the same time period.

Because estimates of NO_2_ were unavailable from CMAQ, we used outputs from the Center for Air, Climate, and Energy Solutions Land Use Regression Models.^43^ These estimates also cover years 2016–2020, are publicly available, and are specifically designed to estimate 2010 tract concentrations. We averaged annual mean NO_2_ tract concentrations from 2016–2020.

To assess tract-level wildfire PM_2.5_, we used the 2016–2023 public estimates by Childs et al.,^44^ which isolate wildfire-specific PM_2.5_ from other PM_2.5_ sources, and accurately capture high-concentration events and daily temporal variation. We averaged daily wildfire PM_2.5_ for each tract from 2016–2023 (versus 2016–2020 for other pollutants) because fire activity has increased substantially since 2020 due to climate change, and because yearly variability is high, making longer averages more representative of wildfire smoke exposure.^10,44^

A small proportion (0.5%, n=411) of census tracts had missing exposure data for one or more pollutants, representing 0.1% of the total contiguous U.S. population. The prevalence of ADL limitations within census tracts missing exposure data was similar to the overall prevalence (14% with ADL limitations among tracts missing exposure data versus 12% overall ADL limitation prevalence).

### Analysis

We conducted a two-part analysis. First, we created choropleth maps to identify census tracts with high ADL limitation prevalence and high air pollution exposure to PM_2.5_, O_3_, NO_2_, and wildfire PM_2.5_. Second, using counts of people with ADL limitations broken down by age, race, and poverty status in each census tract, we calculated the mean, median, standard deviation, and range of exposures to PM_2.5_, O_3_, NO_2_, and wildfire PM_2.5_ among people with ADL limitations across sociodemographic characteristics (age, race, poverty) and by urbanicity and state. These factors predict air pollution exposure, and we aimed to identify highly exposed subgroups within the population with ADL limitations. We made no hypotheses about where high ADL limitation prevalence co-occurs with high exposure, and we hypothesized that racialized people and people with incomes below the federal poverty threshold were more exposed to PM_2.5_ and NO_2_ compared to White people and those above the federal poverty threshold.

As a secondary analysis, we tested for disparities in air pollution exposure among people with ADL limitations versus those without, hypothesizing that people with ADL limitations were more exposed to all four pollutants. We calculated unadjusted mean, median, standard deviation and range of exposures to PM_2.5_, O_3_, NO_2_, and wildfire PM_2.5_ among people with ADL limitations and those without.

### Sensitivity analysis

Since non-ACS estimates of ADL limitation prevalence in prisons and other group quarters vary, are based on small samples, or may not generalize across the country, we repeated all analyses excluding the group quarters population (2.4% of contiguous U.S. population).

## Results

Our analysis included 321 million people living outside group quarters in ~73,000 contiguous U.S. census tracts. 40 million (12%) of those included reported ADL limitations on the 2016-2020 ACS. We estimated 2.5 million people in institutions (68%) had ADL limitations based on reported prevalences.^28–33,35^ People with ADL limitations were distributed throughout the country, and most dense urban centers had areas high ADL limitation prevalence; for example in New York City, certain areas of Manhattan and the Bronx had high prevalence of ADL limitations (**Figure 1**).

**Figure 1:**
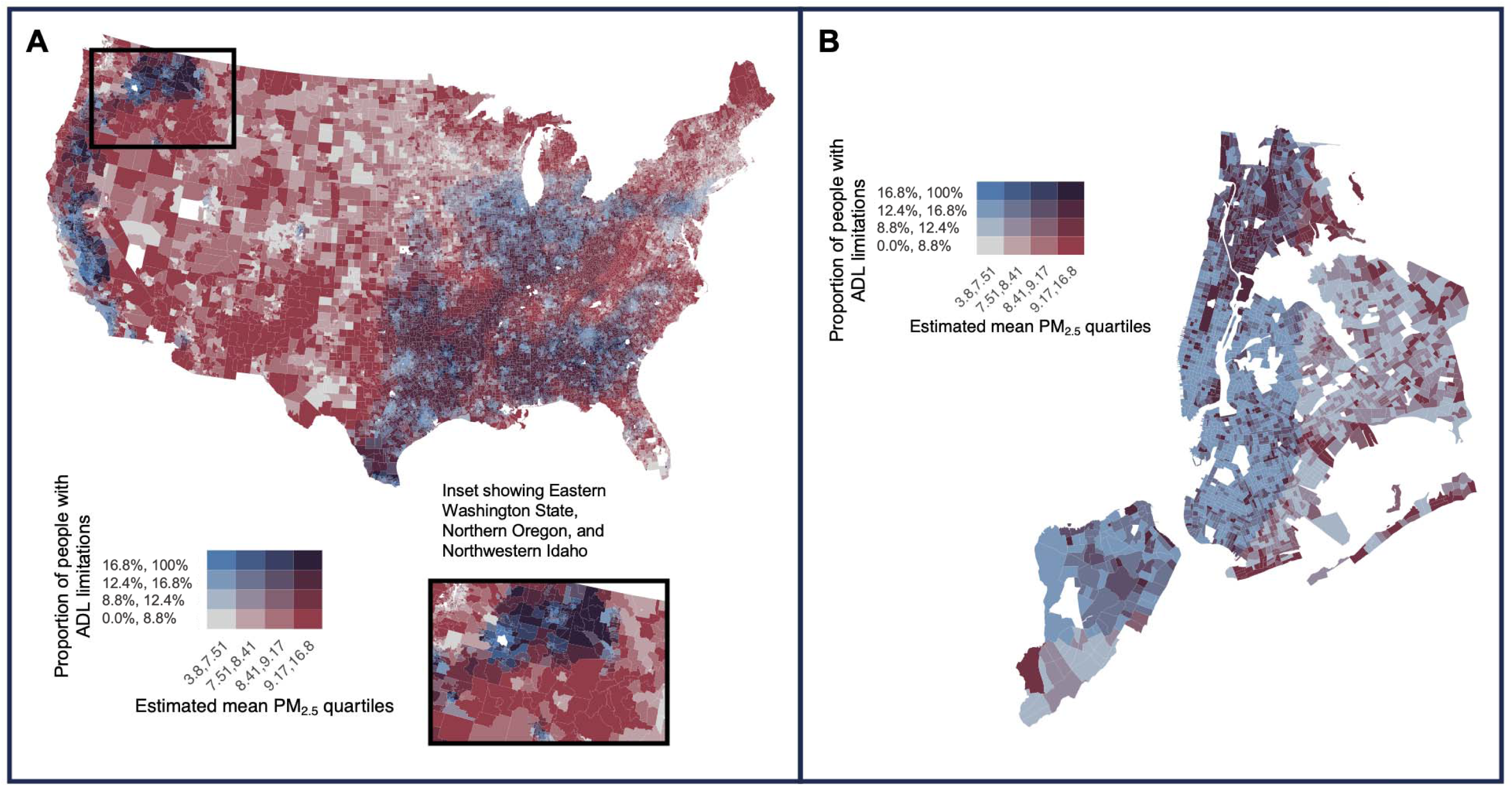
**A**: Co-occurrence of high ADL limitation prevalence and high estimated 2016-2020 mean all-source PM_2.5_ concentration (μg/m^3^) in the contiguous U.S. Map inset shows the area of Eastern Washington State, Northern Oregon, and Northwestern Idaho with high disability prevalence and high all-source PM_2.5_ concentrations. Legend shows quartiles of people with ADL limitations. **Figure 1B:** Co-occurrence of high ADL limitation prevalence and high estimated 2016-2020 mean all-source PM_2.5_ concentrations (μg/m^3^) in New York City. Legend shows quartiles of people with ADL limitations.

Between 2016–2020, for the total population including people in group quarters, the median population-weighted outdoor estimated concentration of PM_2.5_ was 8.43 μg/m^3^ (interquartile range, IQR = 7.54, 9.20), of O_3_ was 37.46 parts per billion by volume (ppbv) (IQR = 36.14, 38.85), and of NO_2_ was 5.43 parts per billion (ppb) (IQR = 3.55, 7.86). Between 2016– 2023, average wildfire PM_2.5_ concentrations were 0.29 μg/m^3^ (IQR = 0.19, 0.45) (**Table 1**).

**Table 1:** Median (IQR) exposure to PM_2.5_, O_3_, NO_2_, and wildfire-generated PM_2.5_ 2016-2020 for United States population by disability status, and by disability status and age, race, poverty, and urbanicity.

| Group | Total N | Median PM<sub>2.5</sub><br>exposure (IQR)<br>in µg/m<sup>3</sup> | Median O<sub>3</sub><br>exposure (IQR) in<br>ppbv | Median NO<sub>2</sub><br>exposure (IQR) in<br>ppb | Median wildfire<br>PM<sub>2.5</sub> exposure<br>(IQR) in µg/m<sup>3</sup> |
| --- | --- | --- | --- | --- | --- |
| Total population | 321,525,041 | 8.43 (7.55, 9.20) | 37.46 (36.14, 38.86) | 5.44 (3.56, 7.87) | 0.29 (0.19, 0.45) |
| Total population with ADL limitations no group quarters | 40,786,461 | 8.38 (7.49, 9.11) | 37.47 (36.22, 38.75) | 5.06 (3.20, 7.51) | 0.28 (0.19, 0.43) |
| Total population no ADL limitations no group quarters | 280,738,580 | 8.44 (7.56, 9.21) | 37.46 (36.13, 38.87) | 5.49 (3.61, 7.92) | 0.29 (0.19, 0.46) |
| Total population including institutions | 329,764,057 | 8.43 (7.54, 9.20) | 37.46 (36.14, 38.85) | 5.43 (3.55, 7.86) | 0.29 (0.19, 0.45) |
| Total population ADL limitations including group quarters | 43,253,294 | 8.37 (7.49, 9.11) | 37.47 (36.22, 38.73) | 5.03 (3.19, 7.47) | 0.28 (0.19, 0.43) |
| Total population no ADL limitations inc group quarters | 286,510,762 | 8.44 (7.55, 9.21) | 37.46 (36.13, 38.87) | 5.49 (3.61, 7.93) | 0.29 (0.19, 0.46) |
| Group quarters population | 3,682,458 | 8.28 (7.40, 8.95) | 37.41 (36.24, 38.60) | 4.26 (2.90, 6.41) | 0.29 (0.19, 0.42) |
| Total with ADL limitations (0-18) | 3,166,556 | 8.44 (7.58, 9.15) | 37.50 (36.30, 38.74) | 5.23 (3.41, 7.55) | 0.29 (0.19, 0.43) |
| Total without ADL limitations (0-18) | 69,996,565 | 8.48 (7.62, 9.24) | 37.51 (36.20, 38.95) | 5.43 (3.61, 7.81) | 0.30 (0.20, 0.46) |
| Total with ADL limitations (18-65) | 20,231,217 | 8.40 (7.53, 9.12) | 37.49 (36.27, 38.72) | 5.15 (3.22, 7.61) | 0.29 (0.19, 0.43) |
| Total without ADL limitations (18-65) | 177,058,479 | 8.46 (7.58, 9.22) | 37.45 (36.11, 38.87) | 5.61 (3.70, 8.09) | 0.29 (0.19, 0.46) |
| Total with ADL limitations (65+) | 17,388,688 | 8.33 (7.44, 9.09) | 37.44 (36.15, 38.77) | 4.92 (3.13, 7.37) | 0.28 (0.19, 0.43) |
| Total without ADL limitations (65+) | 33,683,536 | 8.28 (7.37, 9.08) | 37.39 (36.11, 38.69) | 4.94 (3.23, 7.27) | 0.27 (0.19, 0.43) |
| White with ADL limitations | 30,123,435 | 8.26 (7.37, 9.00) | 37.50 (36.33, 38.74) | 4.57 (2.98, 6.76) | 0.28 (0.19, 0.43) |
| White without ADL limitations | 196,660,632 | 8.33 (7.42, 9.09) | 37.47 (36.24, 38.82) | 4.95 (3.30, 7.13) | 0.29 (0.19, 0.45) |
| Black with ADL limitations | 5,589,694 | 8.66 (8.04, 9.37) | 37.16 (36.02, 38.26) | 6.47 (4.27, 9.14) | 0.27 (0.19, 0.34) |
| Black without ADL limitations | 34,379,889 | 8.62 (7.99, 9.35) | 37.20 (36.00, 38.31) | 6.40 (4.48, 8.86) | 0.26 (0.19, 0.34) |
| American Indian or Alaska Native with ADL limitations | 443,753 | 7.94 (6.35, 8.96) | 38.31 (36.66, 43.81) | 4.01 (2.44, 6.87) | 0.38 (0.25, 0.56) |
| American Indian or Alaska Native without ADL limitations | 2,178,843 | 7.92 (6.22, 9.02) | 38.79 (36.80, 44.98) | 4.26 (2.53, 7.37) | 0.39 (0.26, 0.57) |
| Asian with ADL limitations | 1,323,361 | 8.97 (8.03, 9.98) | 37.35 (35.32, 40.64) | 8.01 (5.86, 11.48) | 0.35 (0.19, 0.68) |
| Asian without ADL limitations | 17,002,053 | 8.85 (8.04, 9.82) | 37.22 (35.35, 39.55) | 7.67 (5.69, 10.91) | 0.33 (0.19, 0.61) |
| Pacific Islander with ADL limitations | 67,357 | 8.56 (7.52, 9.72) | 38.25 (35.29, 43.19) | 7.09 (4.60, 9.92) | 0.35 (0.00, 0.83) |
| Pacific Islander without ADL limitations | 529,742 | 8.60 (7.59, 9.74) | 38.29 (35.18, 43.23) | 7.09 (5.01, 9.77) | 0.40 (0.00, 1.11) |
| Other Race with ADL limitations | 1,508,956 | 8.71 (7.86, 9.95) | 37.79 (35.70, 43.16) | 8.24 (5.53, 13.21) | 0.33 (0.19, 0.55) |
| Other Race without ADL limitations | 15,069,770 | 8.88 (8.01, 10.14) | 37.99 (35.80, 43.01) | 8.44 (5.69, 13.40) | 0.35 (0.21, 0.57) |
| Two or More Races with ADL limitations | 1,729,905 | 8.48 (7.58, 9.28) | 37.58 (36.00, 39.63) | 6.02 (3.91, 8.51) | 0.31 (0.19, 0.51) |
| Two or More Races without ADL limitations | 14,917,651 | 8.56 (7.65, 9.44) | 37.64 (35.92, 39.90) | 6.28 (4.35, 8.81) | 0.32 (0.19, 0.51) |
| Hispanic with ADL limitations | 5,396,728 | 8.63 (7.69, 9.73) | 37.78 (35.54, 42.98) | 7.11 (4.90, 10.85) | 0.33 (0.19, 0.54) |
| Hispanic without ADL limitations | 53,199,763 | 8.76 (7.78, 9.89) | 37.90 (35.73, 42.81) | 7.12 (4.98, 10.79) | 0.33 (0.20, 0.54) |
| ADL limitations below poverty | 8,164,210 | 8.42 (7.56, 9.13) | 37.41 (36.12, 38.62) | 5.43 (3.25, 8.11) | 0.28 (0.19, 0.42) |
| No ADL limitations below poverty | 32,726,712 | 8.53 (7.67, 9.26) | 37.46 (36.05, 38.92) | 5.90 (3.69, 8.57) | 0.30 (0.20, 0.45) |
| ADL limitations above poverty | 32,408,167 | 8.36 (7.48, 9.11) | 37.49 (36.24, 38.79) | 4.98 (3.18, 7.36) | 0.28 (0.19, 0.43) |
| No ADL limitations above poverty | 244,451,152 | 8.43 (7.55, 9.21) | 37.46 (36.14, 38.87) | 5.43 (3.60, 7.84) | 0.29 (0.19, 0.46) |
| Metropolitan with ADL limitations | 34,998,057 | 8.46 (7.59, 9.21) | 37.46 (36.21, 38.82) | 5.63 (3.72, 8.00) | 0.28 (0.19, 0.43) |
| Metropolitan without ADL limitations | 251,595,816 | 8.51 (7.65, 9.29) | 37.46 (36.11, 38.91) | 5.88 (4.04, 8.30) | 0.29 (0.19, 0.46) |
| Large Town with ADL limitations | 3,606,920 | 8.03 (7.02, 8.61) | 37.64 (36.48, 38.62) | 2.89 (2.12, 3.84) | 0.29 (0.22, 0.43) |
| Large Town without ADL limitations | 18,484,475 | 7.99 (6.88, 8.62) | 37.61 (36.42, 38.65) | 2.95 (2.19, 3.94) | 0.30 (0.22, 0.45) |
| Small Town with ADL limitations | 1,293,307 | 7.95 (6.98, 8.53) | 37.35 (36.13, 38.22) | 2.62 (1.99, 3.39) | 0.30 (0.23, 0.41) |
| Small Town without ADL limitations | 6,214,163 | 7.89 (6.91, 8.52) | 37.34 (36.13, 38.25) | 2.72 (2.05, 3.48) | 0.31 (0.23, 0.43) |
| Rural with ADL limitations | 859,424 | 7.40 (6.37, 8.27) | 37.21 (36.08, 38.06) | 2.14 (1.63, 2.68) | 0.29 (0.20, 0.45) |
| Rural without ADL limitations | 4,242,492 | 7.31 (6.29, 8.24) | 37.12 (36.04, 38.04) | 2.25 (1.70, 2.76) | 0.31 (0.21, 0.49) |

### Co-occurrence of high ADL limitation prevalence and high exposure

High estimated outdoor all-source PM_2.5_ exposure co-occurred with high ADL limitation prevalence in California’s Central Valley and urban areas, Washington State and Idaho, and some areas of the Southeast (**Figure 1A**). Many urban areas, where all-source PM_2.5_ exposure was high, had some neighborhoods where exposure co-occurred with high ADL limitation prevalence. For example, in New York City, many areas of Upper Manhattan and the Bronx had both high ADL limitations prevalence and all-source PM_2.5_ exposure (**Figure 1B**). Areas in Eastern Washington State and Northwestern Idaho had both high PM_2.5_ concentrations and high ADL limitation prevalence (**Figure 1A**).

NO_2_ exposure co-occurred with high ADL limitation prevalence in a subset of areas with high street network density or highways, mostly in urban areas (**Supplemental Figure 5**). Ozone exposure co-occurred with high ADL limitation prevalence in the Southwest, as well as neighbourhoods of urban areas with high ozone levels, like Denver, Colorado (**Figure 3**). Wildfire PM_2.5_ exposure co-occurred with high ADL limitation prevalence wherever there was high ADL limitation prevalence in the Pacific Northwest, including Eastern Washington State and Northwestern Idaho (**Supplemental Figure 6**).

**Figure 2:**
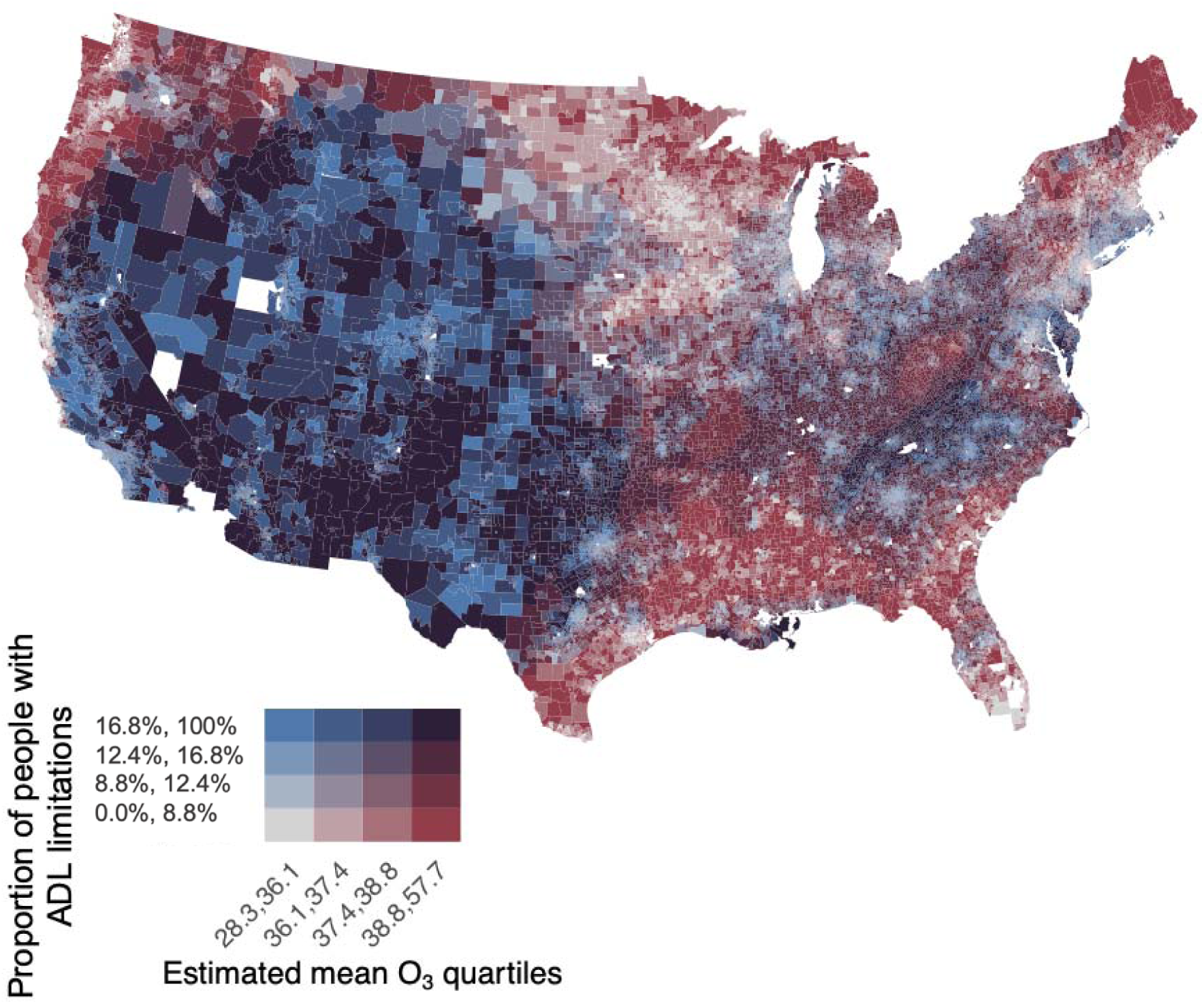
Co-occurrence of high ADL limitation prevalence and high estimated mean 2016-2020 ozone concentrations (ppbv) in the contiguous U.S. Legend shows quartiles of people with ADL limitations.

**Figure 3:**
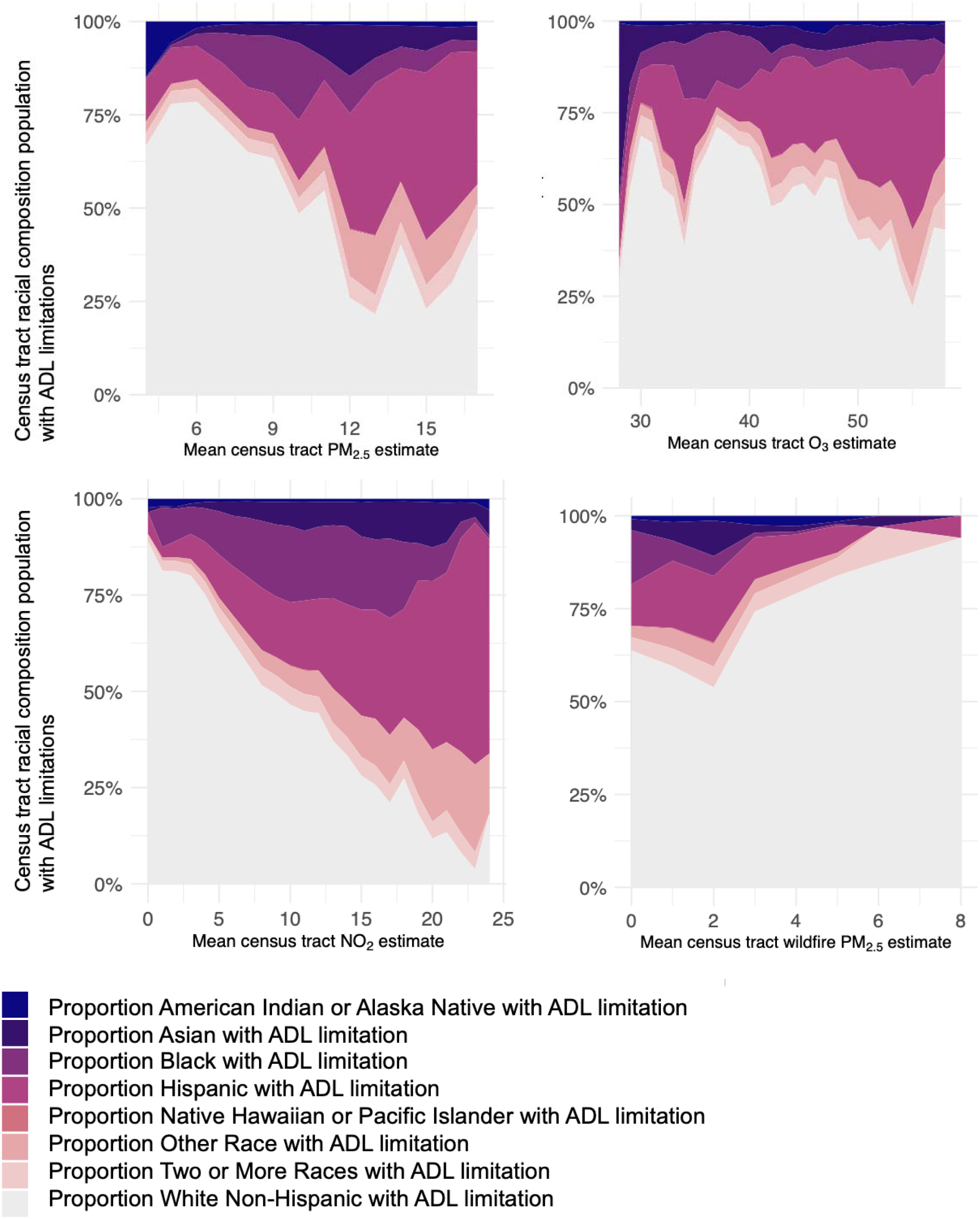
Average census tract-level racial and ethnic composition of people with ADL limitations based on 2016-2020 ACS data in the contiguous U.S. for (a) mean 2016-2020 all-source PM_2.5_ concentration (μg/m^3^); (b) mean 2016-2020 ozone concentration (ppbv); (c) mean 2016-2020 NO_2_ concentration (ppb); and (d) mean 2016-2023 wildfire PM_2.5_ concentration (μg/m^3^).

These patterns did not change when people living in group quarters were excluded from the analysis.

### Exposure within sociodemographic categories of populations with ADL limitations

Among people with ADL limitations, air pollution exposure varied by race, poverty status, and urbanicity. Hispanic people with ADL limitations were disproportionately exposed to all-source PM_2.5_, O_3_, and NO_2_ compared to mean population exposure, while Asian people with ADL limitations were disproportionately exposed to PM_2.5_ and NO_2_. Non-Hispanic White people with ADL limitations were disproportionately exposed to wildfire PM_2.5_ compared to population mean exposure, with higher tract-level proportions of White people associated with higher wildfire PM_2.5_ exposure and lower NO_2_ exposure (**Figure 3**).

Among people with ADL limitations, those with incomes below the federal poverty threshold had higher average PM_2.5_ and NO_2_ exposures compared to those with incomes above the federal poverty threshold, though O_3_ and wildfire PM_2.5_ exposures were similar across poverty levels (**Supplemental Table 1**). Rural versus urban populations with ADL limitations had lower PM_2.5_ and NO_2_ exposure (**Supplemental Tables 1, Supplemental Table 3**), and people age 65+ had lower PM_2.5_ and NO_2_ exposure compared to younger people (**Supplemental Table 1, Supplemental Table 3**).

These patterns did not change when people living in group quarters were excluded from the analysis.

### Differences in exposure by ADL limitations versus no limitations

People with ADL limitations were slightly less exposed to PM_2.5_ and NO_2_ compared to those without, regardless of whether group quarters were included (**Supplemental Figures 1 and 2**). From 2016-2020, the median PM_2.5_ exposure was 8.37 μg/m^3^ (IQR = 7.49, 9.11) for people with ADL limitations versus 8.44 μg/m^3^ (IQR = 7.55, 9.21) for those without (**Table 1**). Median outdoor NO_2_ concentrations were 5.03 ppb (3.19, 7.47) for people with ADL limitations versus 5.49 ppb (IQR = 3.61, 7.93) for those without (**Supplemental Figures 2 and 4**). O_3_ and wildfire PM_2.5_ concentrations were nearly identical between groups, regardless of whether group quarters were included: 37.47 ppb (IQR = 36.22, 38.73) for people with ADL limitations versus 37.46 (IQR = 36.13, 38.87) (**Supplemental Figure 2**) and 0.28 μg/m^3^ (IQR = 0.19, 0.43) for people with ADL limitations versus 0.29 μg/m^3^ (0.19, 0.46).

When investigating exposure among people outside group quarters within categories of age, race, poverty status, urbanicity, and within each contiguous U.S. state, we still found people with ADL limitations to be slightly less or equally exposed to PM_2.5_, O_3_, NO_2_, and wildfire PM_2.5_ compared to those without ADL limitations (**Supplemental Figures 1-4)**.

## Discussion

### Main findings

We examined exposure to PM_2.5_, O_3_, NO_2_ (2016–2020), and wildfire PM_2.5_ (2016–2023) among people with ADL limitations in the contiguous U.S. We identified census tracts with high ADL limitation prevalence and high air pollution exposure, and highly exposed subpopulations. Racial and socioeconomic disparities among people with ADL limitations mirrored those in the general population: racialized and low-income groups were more exposed to air pollution.

Inaccessible healthcare, housing, and transportation can exacerbate air pollution risks for disabled people, as can disability-related health conditions such as chronic obstructive pulmonary disease or asthma. A South Korean study found that short-term air pollution exposure was more strongly associated with hospitalization for cardiovascular causes among people with several types of disability compared to those without.^9^ Another South Korean study found an association between short-term increases in all-source PM_2.5_ and hospitalization for a range of disease categories (e.g., cardiovascular, genitourinary, and respiratory). This association was stronger among people with a range of disabilities versus those without.^18^ Our findings highlight populations with ADL limitations who face high exposure and intersecting social vulnerability, indicating health risk.

### Wildfire PM_2.5_ findings

Disability advocacy organizations have highlighted disabled people’s vulnerability to wildfire PM_2.5_ exposure, especially disabled people with respiratory conditions or mobility limitations.^4,45,46^ Wildfire smoke is unpredictable and can quickly reach harmful levels.^47^ Exclusionary smoke emergency planning leaves disabled people at risk. Emergency alerts are often not provided in accessible formats, such as ASL, captioning, or plain-language text. Shelters may lack wheelchair access, accessible bathrooms, refrigeration and backup power, or accommodations for caregivers and service animals. Emergency response plans frequently assume people can drive or move independently.^48^ Power shutoffs or outages during smoke events also endanger those who use life-sustaining electricity-dependent medical devices such as ventilators or oxygen tanks.^49,50^

Although we found that people with ADL limitations are, on average, similarly exposed to wildfire PM_2.5_ compared to those without ADL limitations, there are areas where high ADL limitation prevalence co-occurs with high smoke exposure such as the Pacific Northwest and California’s Central Valley. These regions are increasingly affected by wildfire smoke due to climate change, and disabled people in these places are at high risk of smoke-related health effects. Interventions should prioritize these high-risk populations.

Although we did not specifically analyze institutionalized populations, wildfire smoke may pose particular dangers in institutions,^14^ which remove residents’ autonomy to respond and whose inadequate or nonexistent responses may further threaten health.^51^

### Exposure differences between those with ADL limitations vs no limitations

In a secondary disparities analysis, we compared air pollution exposure among people with and without ADL limitations. Unexpectedly, people with ADL limitations had slightly lower median exposures to all-source PM_2.5_ and NO_2_, and similar exposures to O_3_ and wildfire PM_2.5_. This pattern held across racial, ethnic, poverty, urbanicity, and regional subgroups.

Based on prior literature and community knowledge of disability and environmental injustice, we did not expect this finding. Environmental injustice has increased disability rates among groups disproportionately exposed to pollution, such as Black and low-income populations.^52,53^ Additionally, disability, race, environmental quality, and income are bi-directionally linked: low-income individuals face higher risk of developing disability due to limited healthcare, unsafe work, poor housing, and environmental exposures, while ableism limits economic opportunities for disabled people.^52,53^ Chakraborty^20^ found that census tracts with higher disability prevalence in people under 65 had higher all-source PM_2.5_ concentrations. Chakraborty defined disability as limitations in ADLs, assessed exposure from 2011-2015, and controlled for race and ethnicity, poverty, renter occupancy, population density, and metropolitan status. In contrast, our study did not test whether disability predicted PM_2.5_ exposure but instead described unadjusted exposure distributions among people with and without ADL limitations.

We offer six possible reasons why our national-scale data may not capture true existing disparities. First, while disability rates are elevated in environmental justice communities, most people with ADL limitations identified by the ACS are White (74%) and many are age 65+ (43%). Therefore, while Black and low-income people are overrepresented among the disabled population, most people with ADL limitations included in this study are not from groups facing the highest pollution exposure. Additionally, White people may be more likely to report ADL limitations and identify as disabled, meaning underreporting in other groups could affect results.^54,55^ Second, higher mortality among those heavily exposed to pollution could create selection bias. People most heavily exposed to pollution earlier in life may be likely to die before reaching old age and developing disability, and therefore may be under-represented among observed survivors in our study. The surviving disabled population may be less exposed. Third, within-census tract variability in pollution exposure was not captured in our data, and could contribute to disparities. Individual-level or census block data may be more accurate. Fourth, some people may relocate to cleaner areas after becoming disabled: health-driven relocation that could lower average exposure among those with ADL limitations. Fifth, while we did not observe differences in pollution exposure by ADL limitations in any age group, much of our study population was 65+. ADL limitations may have different socioeconomic effects in older people compared to younger people and may not lead to losses in income or educational opportunities that may force relocation to more polluted areas. Indeed, older adults and retirees live in more rural places with lower pollution levels compared to younger and working people.^56^ Finally, ADL limitations may not fully capture disability-related marginalization. ACS questions about ADLs were originally designed to assess functioning in older adults. Many people experience disability at some point in their lives, but its impact on socioeconomic status and residence in polluted areas varies widely and is determined by factors such as poverty, racism, and social support. Future studies could consider using dual Medicare–Medicaid eligibility as a proxy for disability-related marginalization. Dual Medicare–Medicaid eligibility often reflects disability and socioeconomic disadvantage, since individuals qualify only if they have significant health needs and low income.

### Interventions to prevent air pollution-related adverse health outcomes in disabled people

Here, we discuss possible interventions for reducing air pollution exposure and harms among disabled people. These interventions should be applied where high air pollution exposure co-occurs with high disability prevalence, as in the regions and subpopulations identified in our results. We underscore that effective interventions require full participation of disabled people in research, design, and implementation---a core principle of disability justice.^5^ Community organizations like Mask Oakland already provide N95 and KN95 masks to unhoused people, many of whom are disabled, in smoke emergencies.^57^ If clean air centers^1^ are accessible, they may be effective in reducing exposure. Providing backup batteries during power outages and shutoffs in programs like PG&E’s backup battery program can protect health during wildfire smoke events by powering medical devices and air filtration. However, these programs have financial and administrative barriers to participation, such as requiring documentation of medical diagnosis. Communication about air pollution emergencies must be delivered in accessible, multimedia formats, and evacuation and sheltering plans must be inclusive. Expanding access to urgent care via telehealth or in-home services can help disabled individuals receive timely treatment without increasing outdoor air pollution exposure. Support and funding for in-home rather than institutional care can ensure autonomy and assistance for disabled people, as institutions have often failed to respond adequately or at all during air pollution emergencies.^14,58^

Additionally, broader environmental justice interventions benefit disabled populations. Measures such as stricter air quality regulations, reducing traffic emissions through electric vehicle incentives, congestion pricing, and urban design to limit air pollution exposure near highways could be beneficial. Policymakers should also reference the fourth and sixth Principles of Environmental Justice from the 1991 First National People of Color Environmental Leadership Summit,^59^ which call for ending the production of air pollution and ensuring universal protection from it.

## Limitations

We could only assess ADL limitation status by race and ethnicity using the available non-mutually exclusive and insufficiently descriptive ACS categories (e.g., Black alone, Non-Hispanic White alone, Asian American alone, Pacific Islander or Native Hawaiian alone, American Indian and Alaska Native alone, Other alone, two or more races, and Hispanic). The ACS only reported poverty status as a binary variable.

Additionally, the ACS defines disability based on limitations in activities of daily living and excludes those living in institutions. This medicalized definition captures only one aspect of disability, excludes disabled people with certain chronic illnesses and mental health conditions, and may not capture marginalization due to disability. Social attitudes, systemic barriers and marginalization, shared culture and experiences, and community make disability a complex experience not fully described by functional limitations. People with ADL limitations do not always describe themselves as disabled.^60^ The ACS also does not disaggregate data describing populations living in group quarters by disability status at the census tract scale, so we relied on national estimates and smaller non-representative surveys of ADL limitation prevalence.

Finally, our study described patterns in air pollution exposure at the national level with census tract scale air pollution estimates. Future studies must account for local variation in both pollution levels and population characteristics. Some pollutants, like ozone and wildfire smoke, affect large areas uniformly, while others—such as traffic-related pollutants—vary significantly at the neighborhood level. Interventions must consider local variation in exposure and community need.

## Conclusion

We identified geographic areas of the contiguous U.S. where high ADL limitation prevalence co-occurred with high air pollution exposure, and described highly exposed and vulnerable racial, ethnic, and socioeconomic subgroups within this population. We observed racial, ethnic, and socioeconomic disparities in exposure within the population with ADL limitations that mirror those observed in overall population. Disabled people are made vulnerable to air pollution-related health effects by social systems. Future research should evaluate interventions to reduce air pollution-related health effects in disabled people, while recognizing that addressing root causes---including structural ableism and environmental injustice---requires transformative change to protect health in this diverse population.

## Supporting information

Supplemental Content

## Data Availability

All data used in this manuscript is publicly available.

## Acknowledgements

We thank Dr. Sunaura Taylor and members of the Disabled Ecologies Lab at UC Berkeley for their transformative feedback on this manuscript. We thank Dr. Barbara Strang for her writing support.

## Footnotes

1 https://ww2.arb.ca.gov/cleanaircenters

## Notes

### Competing Interest Statement

The authors have declared no competing interest.

### Funding Statement

This study was funded by CIHR Doctoral Foreign Study Award (HM), NIEHS P30 P30ES007033 (JAC), and
NIEHS P30 ES009089 (MAK).

