## Supplemental Content for "Air pollution exposure among people with limitations in activities of daily living in the United States"

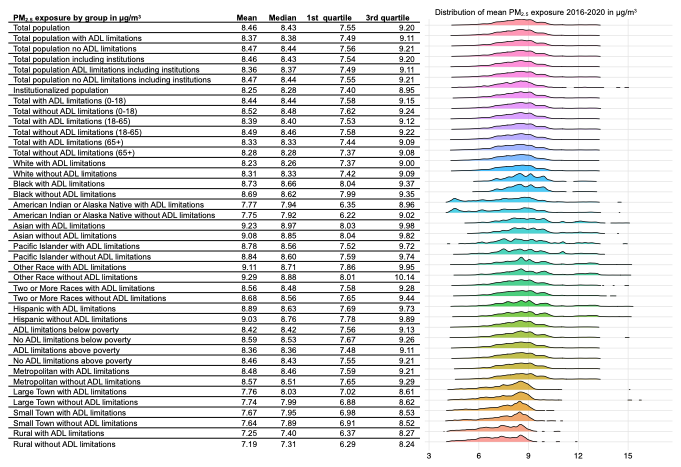

**Supplemental Figure 1**: Mean exposure to PM_2.5_ 2016-2020 for contiguous U.S. population by ADL limitations, and by ADL limitations and age, race, poverty status, and urbanicity as determined from the 2016-2020 American Community Survey. Left panel shows mean, median, and 1^st^ and 3^rd^ quartiles of exposure, while the right panel shows histograms of the distribution of PM_2.5_ for each demographic group.

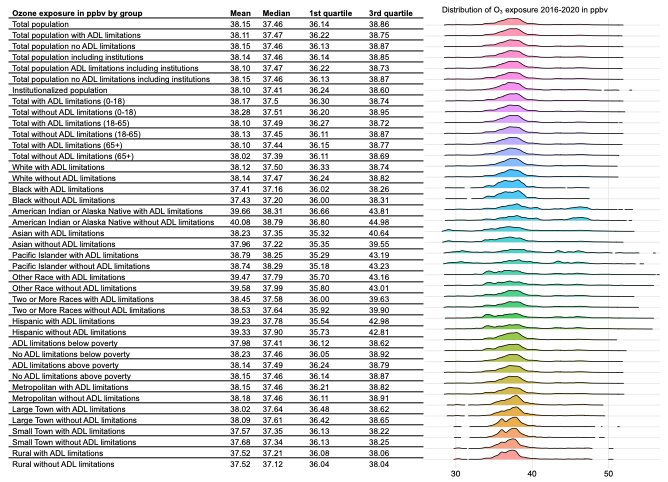

**Supplemental Figure 2**: Mean exposure to ozone 2016-2020 for contiguous U.S. population by ADL limitations, and by ADL limitations and age, race, poverty status, and urbanicity, as determined from the 2016-2020 American Community Survey. Left panel shows mean, median, and 1^st^ and 3^rd^ quartiles of exposure, while the right panel shows histograms of the distribution of ozone for each demographic group.

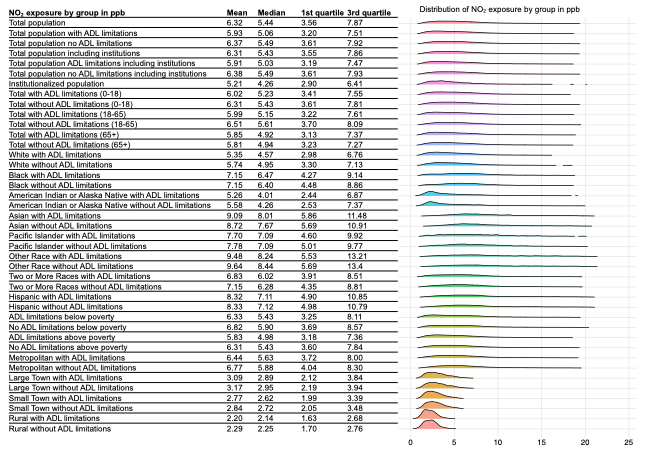

**Supplemental Figure 3**: Mean exposure to NO_2_ 2016-2020 for contiguous U.S. population by ADL limitations, and by ADL limitations and age, race, poverty status, and urbanicity, as determined from the 2016-2020 American Community Survey. Left panel shows mean, median, and 1^st^ and 3^rd^ quartiles of exposure, while the right panel shows histograms of the distribution of NO_2_ for each demographic group.

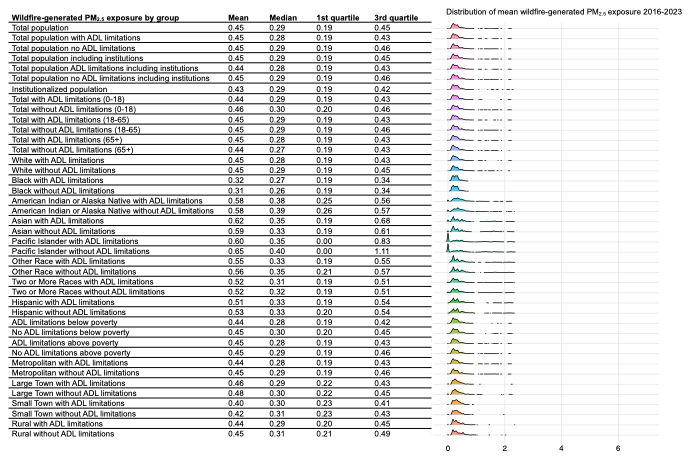

**Supplemental Figure 4**: Mean exposure to wildfire-generated PM_2.5_ 2016-2023 for contiguous U.S. population by ADL limitations, and by ADL limitations and age, race, poverty status, and urbanicity, as determined from the 2016-2020 American Community Survey. Left panel shows mean, median, and 1^st^ and 3^rd^ quartiles of exposure, while the right panel shows histograms of the distribution of wildfire-generated PM_2.5_ for each demographic group.

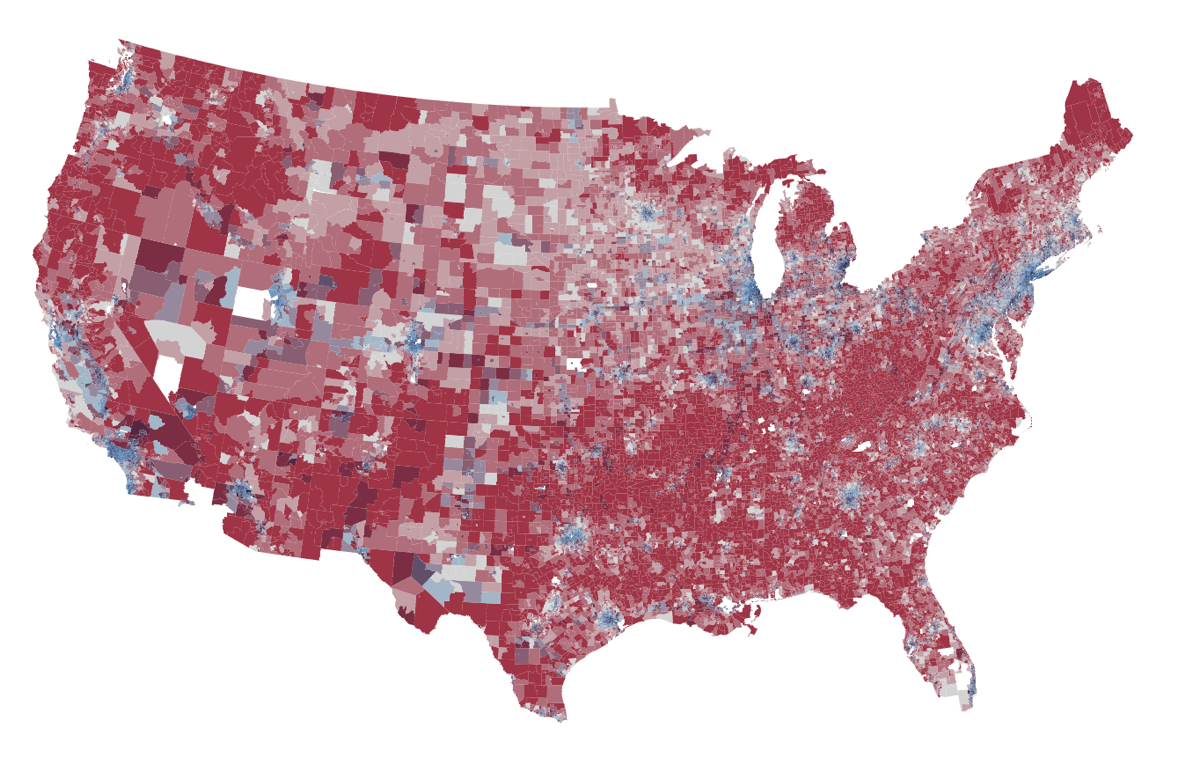

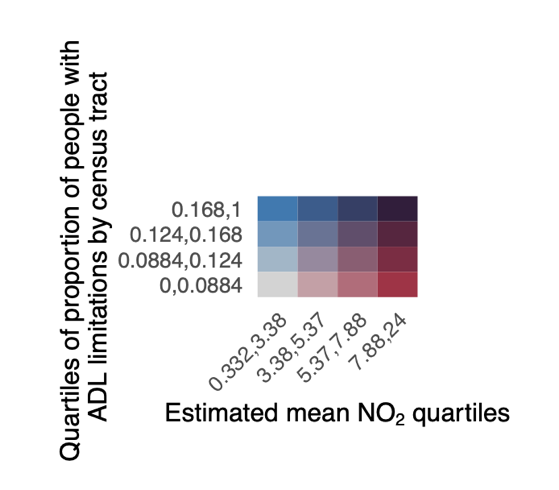

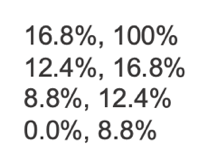

**Supplemental** **Figure 5**: Co-occurrence of high ADL limitation prevalence and high estimated mean 2016-2020 NO_2_ concentration in the contiguous U.S.

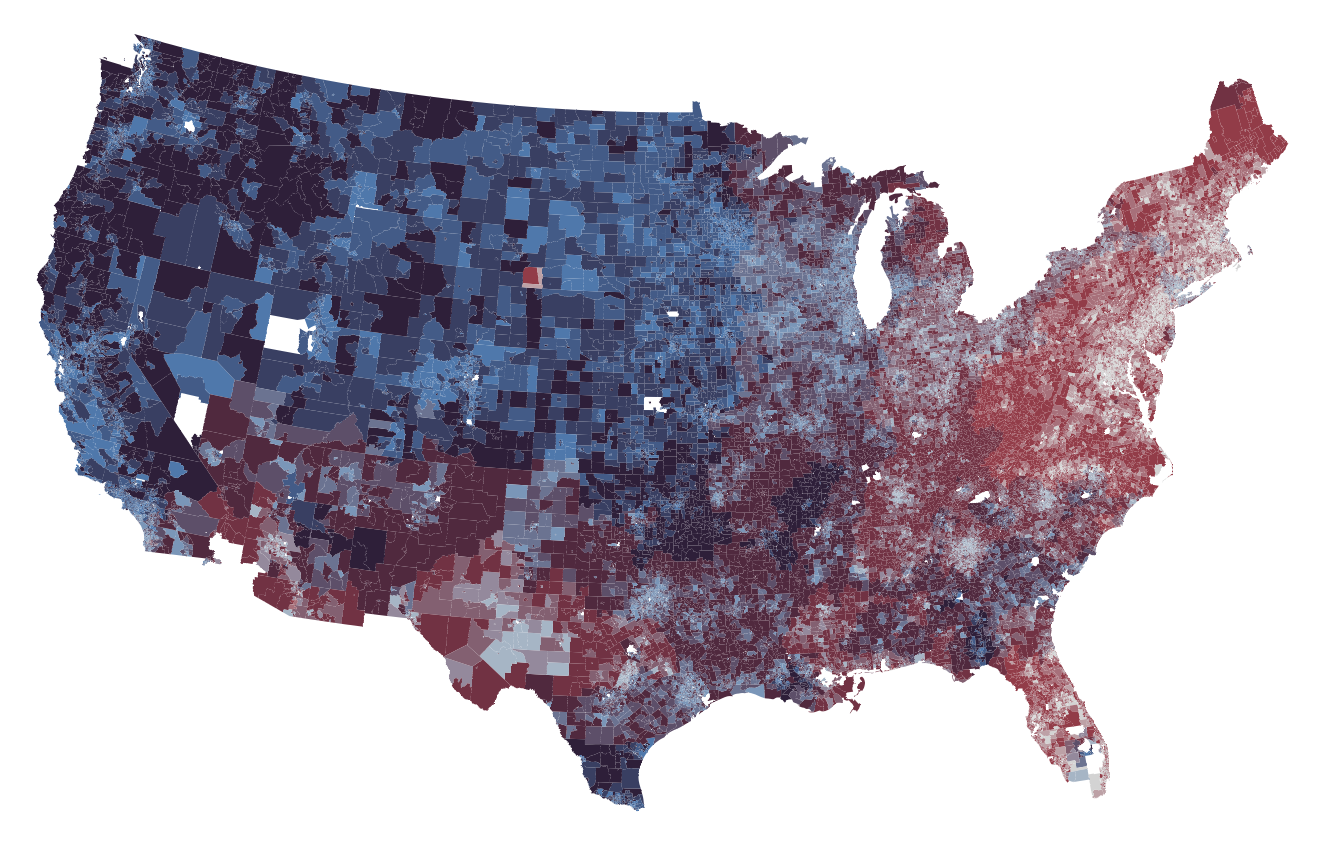

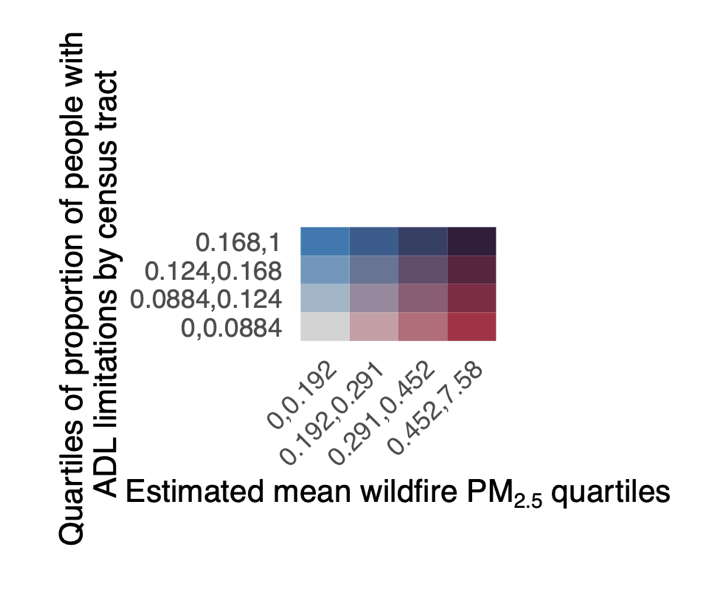

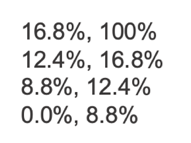

**Supplemental** **Figure 6**: Co-occurrence of high ADL limitation prevalence and high estimated mean 2016-2020 wildfire PM_2.5_ concentration in the contiguous U.S.

**Supplemental Table 1**: Exposure to PM_2.5_, O_3_, NO_2_, and wildfire-generated PM_2.5_ by ADL limitation status and poverty status.

| **Pollutant** | **Demographic group** | **Mean exposure** | **Standard deviation** | **Years** |
| --- | --- | --- | --- | --- |
| PM_2.5_ | No ADL limitation above poverty | 8.36 µg/m^3^ | 1.48 | 2016-2020 |
| PM_2.5_ | ADL limitation above poverty | 8.47 µg/m^3^ | 1.50 | 2016-2020 |
| PM_2.5_ | No ADL limitation below poverty | 8.42 µg/m^3^ | 1.49 | 2016-2020 |
| PM_2.5_ | ADL limitation below poverty | 8.59 µg/m^3^ | 1.59 | 2016-2020 |
| O_3_ | No ADL limitation above poverty | 38.16 ppbv | 3.91 | 2016-2020 |
| O_3_ | ADL limitation above poverty | 38.16 ppbv | 4.05 | 2016-2020 |
| O_3_ | No ADL limitation below poverty | 37.99 ppbv | 3.83 | 2016-2020 |
| O_3_ | ADL limitation below poverty | 38.26 ppbv | 4.15 | 2016-2020 |
| NO_2_ | No ADL limitation above poverty | 5.83 ppb | 3.65 | 2016-2020 |
| NO_2_ | ADL limitation above poverty | 6.31 ppb | 3.80 | 2016-2020 |
| NO_2_ | No ADL limitation below poverty | 6.33 ppb | 4.05 | 2016-2020 |
| NO_2_ | ADL limitation below poverty | 6.82 ppb | 4.22 | 2016-2020 |
| Wildfire-generated PM_2.5_ | ADL limitation above poverty | 0.45 µg/m^3^ | 0.53 | 2016-2023 |
| Wildfire-generated PM_2.5_ | No ADL limitation above poverty | 0.46 µg/m^3^ | 0.51 | 2016-2023 |
| Wildfire-generated PM_2.5_ | ADL limitation below poverty | 0.44 µg/m^3^ | 0.52 | 2016-2023 |
| Wildfire-generated PM_2.5_ | No ADL limitation below poverty | 0.46 µg/m^3^ | 0.52 | 2016-2023 |

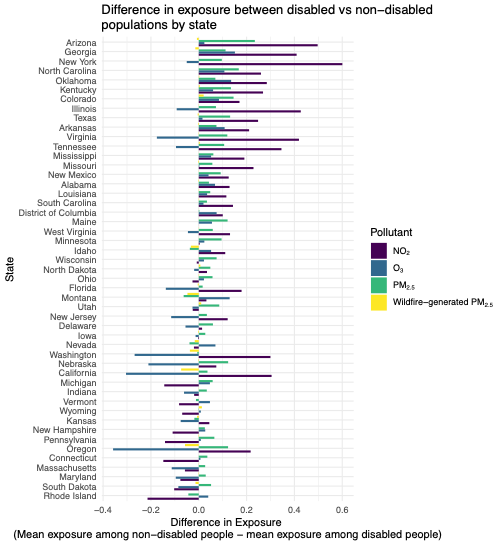

**Supplemental Figure 7**: Differences in mean exposure to all-source PM_2.5_ (µg/m^3^), NO_2_ (ppb), ozone (ppbv) (2016-2020), and wildfire-generated PM_2.5_ (µg/m^3^) (2016-2023) for people with ADL limitations versus no ADL limitations in the contiguous U.S. Difference in exposure is *{mean exposure among people without ADL limitations – mean exposure among people with ADL limitations}*.
